# Exploring intimate partner interference in abortion decisions among people capable of pregnancy in the U.S.: A scoping review protocol

**DOI:** 10.1101/2025.04.26.25326334

**Authors:** Jessica L. Dozier, Karen Trister Grace, Kathryn Vanderboll, Gabrielle Gippert, Vanessa K. Dalton, Yasamin Kusunoki

**Affiliations:** University of Michigan School of Nursing, Department of Systems, Populations and Leadership, Ann Arbor, MI, USA; George Mason University, School of Nursing, Fairfax, VA, USA; University of Michigan Taubman Health Sciences Library, Ann Arbor, MI, USA; University of Michigan College of Literature, Science, and the Arts, Ann Arbor, MI, USA; University of Michigan Medical School, Department of Obstetrics and Gynecology, Ann Arbor, MI, USA

**Keywords:** intimate partner violence, public health, pregnancy coercion, reproductive control, abortion, gender-based violence, abuse, reproductive autonomy

## Abstract

**Background:** Reproductive coercion and abuse (RCA) is a type of intimate partner violence characterized by the use of violence, threats, or manipulation to control an individual’s reproductive health decisions. A key form, abortion coercion, involves compelling an individual capable of pregnancy to have or not have an abortion or limiting their access to abortion care. Understanding intimate partner-perpetrated abortion coercion is increasingly urgent following the U.S. Supreme Court’s 2022 decision to overturn *Roe v. Wade*, which has reshaped reproductive rights and access.

**Objective:** This review aims to systematically examine the available evidence on abortion coercion in the U.S., broadly and in the context of restrictive abortion policies, to understand the prevalence, experiences, and impact of abortion coercion, identify research gaps, and propose recommendations for future research and policy.

**Methods:** We will follow the Joanna Briggs Institute (JBI) Methodology for scoping reviews and adhere to guidelines outlined in the Preferred Reporting of Items for Systematic Review and Meta-Analyses—Scoping Review extension (PRISMA-ScR). Eligible studies will include original research from peer-reviewed articles and grey literature published in English between January 1, 2005 and April 1, 2025. Six databases—Scopus, PubMed, CINAHL, PsycInfo, GenderWatch, and Women’s Studies International—will be searched by a research librarian. Two independent reviewers will assess studies for eligibility; a third reviewer will resolve conflicts. Data will be extracted using a standard form and analyzed using descriptive statistics and qualitative content analysis. Results will be presented in summary tables and visual presentations such as a heat map.

## 1. Introduction

### 1.1 Rationale

Reproductive coercion and abuse (RCA), a type of intimate partner violence (IPV), is increasingly recognized as a significant public health problem worldwide.(1–3) RCA encompasses behaviors by an intimate partner that undermine the autonomy of cisgender women, transgender men, and gender non-binary individuals with the capacity for pregnancy in making decisions about their reproductive health, by exerting control over pregnancy and pregnancy outcomes.(3,4) These behaviors include pregnancy-promoting tactics, such as threats, violence, or manipulation to cause or sustain a pregnancy, as well as pregnancy-preventing behaviors, such as coercing abortion or using threats or violence to prevent conception (1,5). While RCA can impact anyone capable of becoming pregnant, existing research primarily focuses on its perpetration by male intimate partners against cisgender women.(1) In the United States, population-based data indicate that 8.4% of cisgender women experience RCA in their lifetime(6). Clinic-based studies suggest an even higher prevalence, with one in four women attending family planning reporting experiences of RCA.(7) Moreover, significant racial disparities exist, with non-Hispanic Black, multiracial, and Hispanic women experiencing RCA at higher rates than non-Hispanic white women.(6,7)

Abortion coercion is an understudied form of RCA, in which an intimate partner exerts control over an individual’s decision to terminate or continue a pregnancy. This RCA can take multiple forms, including physical violence, manipulation, or blocking access to abortion services.(2) Early research has documented women’s experiences of partner-imposed pressure to resolve their pregnancies in the way their partners want, finding that some who sought to terminate a pregnancy faced guilt, financial control, sabotaged appointments, or even threats of harm.(8) Others who wanted to continue a pregnancy reported being pressured or threatened into terminating it.(8,9) Restrictions on abortion access may further limit reproductive autonomy for those experiencing RCA, making it even more difficult to act against a partner’s control. The urgency of understanding and addressing abortion coercion has grown in the wake of the June 2022 *Dobbs v. Jackson Women’s Health Organization* decision, which overturned *Roe v. Wade* and reshaped abortion rights and access in the U.S.

The *Dobbs* decision enabled a wave of restrictive state-level abortion policies that have exacerbated barriers to care, particularly for those already facing structural inequities. The National Domestic Violence Hotline (NDVH) reports that since *Dobbs,* the number of callers reporting RCA has doubled.(9) In a 2024 study, NDVH found that 13% of respondents were pressured or forced to terminate a pregnancy and 9% faced threats or violence related to abortion decisions.(9) Some reported their partners threatened to involve law enforcement or take legal action against them for considering or obtaining an abortion.(9) Others feared that their abusive partner discovering their abortion could result in arrest or prosecution.(9) These findings underscore the growing risks faced by individuals seeking abortion care in the post-Dobbs era. As legal and social penalties for abortion increase, so do opportunities for abusive partners to exert control. There is an urgent need to systematically synthesize this and other existing research across diverse study designs, populations, and disciplines to better understand how abortion coercion operates in the changing U.S. abortion policy landscape and inform future directions.

While prior reviews have examined RCA broadly (1,10–14) and highlighted the positive role that supportive partners can play in abortion decision-making(15), no review has focused specifically on abortion coercion and examined how policy changes, such as the *Dobbs* decision, may shape coercive and abusive relationship dynamics related to abortion. Existing reviews (1,10–14) rely on narrow search terms that may not capture all aspects of abortion coercion, particularly when studies do not explicitly label these behaviors as “abortion coercion” or “reproductive coercion.” This limitation may partially explain the dearth of literature(1,11) identified on abortion coercion. Additionally, because existing reviews on RCA predate the *Dobbs* decision, they cannot account for the contemporary sociopolitical landscape and its impact on intimate partner interference in abortion decision-making and access.

Understanding people’s experiences with abortion coercion, along with its associated health outcomes and impacts on reproductive autonomy, is essential for guiding future research, policy, and interventions. A scoping review is the most suitable method for exploring this topic, as it allows us to explore the breadth of literature on abortion coercion in an exploratory and descriptive manner. A preliminary search of Scopus, Joanna Briggs Institute (JBI) Evidence Synthesis, PubMed, and Open Science Framework was conducted and no current or underway systematic reviews or scoping reviews on abortion coercion in the U.S. were identified.

While Pike’s 2023 narrative review brings important attention to coerced abortion as a neglected aspect of reproductive coercion, it differs substantially from our proposed scoping review in focus and methodology. Pike provides a global, conceptual overview that includes coercion from multiple sources—such as family members, healthcare providers, and state actors—and draws heavily on opinion, anecdotal reports, and policy critiques. In contrast, our scoping review is limited to partner-perpetrated abortion coercion in the U.S., with a specific emphasis on empirical studies that examine interpersonal dynamics and partner interference in abortion decision-making.

Additionally, our review is the first to explore how recent shifts in U.S. abortion policy, including the *Dobbs* decision, may shape these dynamics. By using broad, inclusive search terms and a systematic approach aligned with JBI methodology, our review seeks to capture a more comprehensive body of literature and map the current evidence base to inform research in this evolving landscape.

Thus, our primary objective is to map the landscape of research on intimate partner-perpetrated abortion coercion in the U.S., both broadly and in the context of restrictive abortion policies, while identifying key gaps that require further investigation.

### 1.2 Research questions

The scoping review will address the following research questions:

*What is known about intimate partner-perpetrated abortion coercion—intimate partner interference in abortion decisions—among people capable of pregnancy in the U.S.?*

A sub-question explores abortion coercion in the context of the changing U.S. abortion policy landscape: *What is known about the relationship between U.S. abortion policy, particularly following the June 2022 Dobbs decision, and intimate partner-perpetrated abortion coercion?*

## 2. Materials and methods

Protocol development involved a multidisciplinary team with expertise in sexual and reproductive health, including reproductive coercion and abortion (JLD, KTG, VKD, YK) and literature synthesis methodology (KV).

The review will be conducted following the JBI Methodology for Scoping Reviews(16,17) and reported in compliance with the Preferred Reporting Items for Systematic Reviews and Meta-Analyses (PRISMA) extension for Scoping Reviews (PRISMA-ScR).(18) Data extraction will occur in Summer 2025 and results are expected in 2026.

Institutional review board approval was not required for this study because human subjects were not involved.

### 2.1 Eligibility criteria

The inclusion criteria are guided by the JBI Population-Concept-Context (PCC) framework.

#### Population

The scope of this review will be limited to studies with participants capable of pregnancy—including cisgender women, transgender men, and non-binary individuals aged 15-49—as well as studies with their current or former intimate partners who have interfered in their abortion decisions. There will be no restrictions on the age or gender of perpetrators. Because RCA can be perpetrated by any individual regardless of sex assigned at birth or sexual orientation, studies of intimate partners of any age and gender will be eligible for inclusion. Additionally, studies reporting on healthcare providers’ and practitioners’ experiences with patients who have experienced abortion coercion will also be included. Reported results will use “women” when reporting on the stated focus and results of prior research. Animal studies will be excluded.

#### Concept

Studies will be considered for inclusion if either of the two abortion coercion research questions are addressed. “Abortion coercion” is defined as any behavior by a current or former intimate partner intended to control the individual’s abortion decisions. This may include, but is not limited to, actions such as forcing a pregnant partner to terminate a wanted pregnancy, using violence or threats to control abortion decisions, or obstructing access to abortion services by a partner. The review will include studies that address the influence of intimate partners on the decision-making process related to abortion, access to abortion services, and post-abortion experiences. Since varying terminology is used in the literature to describe behaviors that may align with abortion coercion, studies are not required to use specific terms like “abortion coercion” or “reproductive coercion.” As long as the behaviors described meet our definition of abortion coercion, the study will be considered for inclusion.

To be eligible for inclusion, studies must explicitly involve partner interference or control over abortion decisions. For example, studies that mention IPV as a reason for seeking abortion but do not describe how an abusive partner directly influenced the decision to have (or not have) an abortion, or other related aspects of abortion decisions, will be excluded. Additionally, studies focusing on coercion by healthcare providers or those studies solely discussing abortion restrictive policy as a form of structural RCA, without addressing interpersonal dynamics of abortion coercion, will be excluded.

Lastly, studies will be included if they discuss physical violence with the intent to cause a miscarriage, but only if accompanied by a partner’s motivation to force the end of the pregnancy after the pregnant person has refused an abortion. This ensures the focus remains on coercion and violence within abortion decision-making rather than broadening to all forms of IPV.

#### Context

The review will be limited geographically to the United States. Original research published on or between January 1, 2005, and April 1, 2025, will be eligible for inclusion. The review begins in 2005 to capture early research on coercion in intimate relationships and reproductive outcomes, including studies that may have reported on intimate partner interference in abortion without using specific terminology related to abortion coercion or RCA.(19) This timeframe also captures a period of increasing scholarly attention to RCA as a distinct form of IPV.(1) It will also capture research conducted in the context of significant shifts in abortion access, policy, and public discourse in the U.S., including the rise of state-level abortion restrictions, the expansion of medication abortion access, and the 2022 reversal of *Roe v. Wade*.

#### Types of Sources

This scoping review will include original research published in peer-reviewed and grey literature. Including grey literature will capture emerging research, unpublished findings, and diverse perspectives that may not be available in traditional academic publishing.

Eligible studies may use quantitative, qualitative, and mixed-methods designs, including both primary and secondary data analyses. Grey literature sources reporting original research, such as preprints from recognized repositories, white papers, theses/dissertations, and organization reports, will be included. Reviews, conference proceedings, editorials, and non-research-based reports will be excluded. Only English-language studies will be included due to the language limitations of the authors.

### 2.2 Search Strategy

KV, a research librarian, developed and refined the search strategy in consultation with the other authors. The complete search strategy for one database (Scopus) is included in S1 Appendix.

We first conducted a preliminary search on Scopus (via Scopus.com) to identify a list of sentinel articles(20–22,8,23,24) that should appear in the search. Citation tracking in Scopus led to the identification of an additional article.(25) Using titles, abstracts, and indexing terms from these articles, we compiled a working list of relevant search terms, including “reproductive coercion” and “pregnancy coercion.” We also reviewed Medical Subject Headings (MeSH), keyword, and index terms from sentinel articles to refine our search terms.

We then conducted a second search using these keywords and index terms across five additional databases: PubMed (via PubMed.gov), CINAHL (via EBSCOHost), PsychInfo, GenderWatch (via ProQuest), and Women’s Studies International (via EBSCOHost).

The initial search strategy was piloted to assess keyword relevance across databases. To validate it, we tested our search strategy against the set of sentinel articles. No geographical, date, or language limits will be applied in the search strategy. Potentially relevant grey literature will be identified through searches in ProQuest Dissertations & Theses Global and Policy Commons. Additional hand searches will be conducted on the websites of the National Domestic Violence Hotline (thehotline.org), Futures Without Violence (futureswithoutviolence.org), and IfWhenHow (ifwhenhow.org).

### 2.3 Source selection and data extraction

#### Initial pilot testing of inclusion criteria and search strategy

During protocol development, two independent reviewers, JLD and GG, screened the titles and abstracts of the first 30 citations obtained from the Scopus search. This process helped authors refine screening protocols and identified any discrepancies in the reviewers’ understanding of inclusion criteria and how to apply them during screening. Specifically, this process helped clarify that the initial title and abstract search will be based solely on these components, not considering any prior knowledge of a data source. When uncertain, we will mark data sources as “maybe” during Covidence screening and proceed to full-text review. This approach enhances reproducibility by accounting for varying levels of background knowledge of the literature among future reviewers. We also determined that at title/abstract screening, all articles on reproductive coercion in U.S. settings published between 2005 and 2025 would automatically proceed to full-text review to ensure we do not miss any results on abortion coercion, whether reported as a secondary finding or without using the term “abortion coercion.”

#### Source selection process

All identified citations will be collated and uploaded into the Covidence software and duplicates will be removed. Reviewers will then screen titles and abstracts against established inclusion criteria. For potentially relevant citations, full texts will be retrieved and imported into Covidence for further evaluation. Two independent reviewers will assess the full text of these citations against the inclusion criteria, documenting conflicts and resolving them through discussion. If consensus is not achieved, a third reviewer will make the final decision. The scoping review report will document and report reasons for excluding citations. Additionally, both forward and backward citation searching of included studies will be conducted via the online tool CitationChaser, which searches the Lens.org database. Articles identified via citation searching will be imported into Covidence and screened for eligibility.

#### Data Extraction

The data extraction form (S2 Appendix) organizes details about participants, concepts, contexts, study methods, and key findings relevant to the review questions. It will be refined iteratively as new relevant data items are identified during data extraction. The final review report will document any additional items not listed in the S2 Appendix, along with justification for the changes.

Reviewers will pilot test the data extraction form (S2 Appendix) and accompanying guidance sheet (S3 Appendix) during the data extraction stage to ensure their comprehensibility and applicability. Two reviewers will independently pilot-test the form on at least one of each type of included full-text evidence source, such as a peer-reviewed article and an NGO report. Reviewers will assess the completeness, clarity, and efficiency of the data extraction form (S2 Appendix) and ensure the accompanying guidance (S3 Appendix) is clear.(26) Additionally, they will document the time required for extraction to inform time allocation for a full review.

After pilot testing of the data extraction form, one reviewer will independently extract data from all the remaining included studies. A second reviewer will review all extracted data for accuracy and completeness. Discrepancies will be resolved through discussion, with a third reviewer making the final decision if necessary.

### 2.4 Data analysis and Presentation

Descriptive statistics (percentages/proportions) will be conducted in Microsoft Excel. Frequency counts will summarize data extraction items, including the number of studies by design (quantitative, qualitative, mixed, or multi-method), location, and study population (e.g., survivors or perpetrators of abortion coercion). We will also determine the number of studies that report abortion coercion aimed at forcing an individual to terminate their pregnancy versus interfering with a wanted abortion. In addition, we will create bar graphs showing the number of sources published by year to descriptively compare the number of sources published each year before and after *Dobbs*.

Following JBI scoping review guidance, a basic qualitative content analysis will also be conducted, using open coding to categorize key concepts across all study designs. We will follow the inductive content analysis process outlined by Pollock et al. (2024), which involves immersion in the data, inductive extraction, open coding, development of a coding framework, categorization, and organization of findings.

The data will be presented in tables that summarize the extracted information. Visual presentations, such as a heat map showing the distribution of included studies across U.S. states or a waffle chart illustrating the methodology used (i.e., quantitative, qualitative, mixed, or multi-methods), will accompany the narrative to describe the results.

Frequent team check-ins will occur during the extraction and analysis phases to discuss the process, address challenges, and review any modifications to the data extraction form or result tables. The rationale for any deviations from this protocol will be documented in the final report.

## 3. Discussion

This scoping review protocol will map all original research on abortion coercion in the U.S. in the last 20 years, which can inform research and policy recommendations. A key strength of this protocol is the inclusion of grey literature, which will capture timely work not typically published in academic journals, such as research from organizations like the National Domestic Violence Hotline, which works directly with individuals experiencing abortion coercion. Additionally, the review will encompass all individuals capable of becoming pregnant—including cisgender women, transgender men, and non-binary people—as well as intimate partners of any gender, ensuring a broad and inclusive perspective on abortion coercion. As is common in scoping reviews,(16) the methodological quality of included studies will not be systematically assessed, as the primary objective is to map the scope of existing research rather than evaluate the risk of bias. Despite this limitation, this scoping review will make an important and timely contribution to the literature. Given the relevance of findings to diverse audiences, results will be shared in a public-facing repository in addition to being submitted for peer-reviewed publication.

## Supporting information

S1 Appendix Search Strategy

S2 Appendix Data Extraction Form

S3 Appendix Data Extraction Guide

S4 Appendix PRISMA-ScR Checklist

## Data Availability

No datasets were generated or analysed during the current study. All relevant data from this study will be made available upon study completion.

## Supporting information captions S1 Appendix. Search Strategy

S2 Appendix. Draft Data Extraction Form

S3 Appendix. Extraction guidance sheet for the scoping review

S4 Appendix. Preferred Reporting Items for Systematic reviews and Meta-Analyses extension for Scoping Reviews (PRISMA-ScR) Checklist

