## Supplementary material for "Exploring intimate partner interference in abortion decisions among people capable of pregnancy in the U.S.: A scoping review protocol": S1 Appendix Search Strategy

**Scopus (via Scopus.com)**

TITLE-ABS-KEY((abort* OR (terminat* AND pregnan*)) AND ("reproductive coercion" OR "reproductive control")) OR TITLE-ABS-KEY (("Intimate partner violence" OR "domestic violence" OR "domestic abuse" OR "spousal abuse" OR "spouse abuse" OR "spousal violence" OR "marital violence" OR "marital abuse" OR "relationship abuse" OR "relationship violence" OR "wife abuse" OR abusive OR "dating violence" OR "interpersonal violence" OR "gender based violence" OR "partner abuse" OR "sexual violence" OR "sexual abuse" OR "emotional abuse" OR "emotional violence" OR "psychological violence" OR "psychological abuse" OR "physical abuse" OR "physical violence" OR "battered women" OR "battered female*" OR "wife beating" OR coerc*) AND (abort* OR (terminat* AND pregnan*) OR "reproductive coercion" OR  "reproductive control" OR "pregnancy coercion" OR "reproductive autonomy" OR "reproductive sabotage" OR "coerced pregnancy" OR "coerced reproduction" OR (promot* W/2 pregnan*)) AND (partner OR partners OR couple OR couples OR husband OR husbands OR wife OR wives OR boyfriend* OR girlfriend* OR spous* OR married))
