## Supplementary material for "Exploring intimate partner interference in abortion decisions among people capable of pregnancy in the U.S.: A scoping review protocol": S2 Appendix Data Extraction Form

**S2 Appendix: Draft Data Extraction Form**

| **Source** | | | |  |  |  |  |  |  |  | **Results** | | |  |  |
| --- | --- | --- | --- | --- | --- | --- | --- | --- | --- | --- | --- | --- | --- | --- | --- |
| **Extractor/**  **Reviewer initials** | **Author (Year)** | **Publication** | **Type of Evidence Source** | **Study Design** | **Objective** | **Population** | **Sample Size** | **Setting** | **Data Collection Methods** | **Concept—nature of abortion coercion** | **Key findings** | **Perpetrator(s)** | **Policy Context** | **Research/**  **Policy Recommendations** | **Gaps/**  **Areas of Further Research** |
