## Supplementary material for "Exploring intimate partner interference in abortion decisions among people capable of pregnancy in the U.S.: A scoping review protocol": S3 Appendix Data Extraction Guide

**S3 Appendix: Extraction guidance sheet for the scoping review** ^1^

| **Extractor/Reviewer Initials** | Initials of researcher conducting initial data extraction and the researcher reviewing all of the extracted data to ensure accuracy and completeness. |
| --- | --- |
| **Author (Year)** | E.g., Davis (2024); Davis *et al.* (for more than two authors) |
| **Publication** | Where was the article published? e.g., peer-reviewed article published in *PLOS ONE, Contraception, or Journal of Midwifery and Women’s Health*). If the source is from the grey literature, indicate where it was published (e.g., report published on the American College of Obstetrics and Gynecology organization website or dissertation published on an institutional repository) |
| **Type of evidence source (primary research/grey literature)** | - Grey literature: e.g., theses and dissertations, preprints, white papers, working papers, government/NGO reports - Primary research: peer-reviewed articles |
| **Objective** | Description of the study objective(s) |
| **Study design** | E.g., randomized control trial (RCT), cohort study, qualitative study, cross-sectional study, case-control study, mixed or multi-methods |
| **Population** | Population studied (e.g., women seeking abortion, general population of women 18-49, men who have perpetrated reproductive coercion, etc.), age, race/ethnicity, other relevant demographic characteristics |
| **Data collection methods** | Interviews, surveys, etc. |
| **Concept—nature of abortion coercion** | Details about the nature of the abortion coercion, including whether it involved pressure to have an abortion or pressure to continue a pregnancy. How is abortion coercion defined, if at all? |
| **Key Findings** | - Prevalence/Incidence rates (if applicable) - Specific abortion coercion behaviors or tactics identified (e.g., emotional manipulation, threats, physical violence, financial control) - Timing of coercion: before pregnancy, during pregnancy, or when seeking abortion services - Key quotes (If qualitative study) - Was abortion coercion a primary or an ancillary finding? |
| **Perpetrator(s)** | Current/former intimate partner, age, relationship type/length |
| **Policy Context** | Does the study discuss the impact of U.S. abortion policy on abortion coercion? Does the study examine abortion coercion in the context of the post-Dobbs decision landscape? Describe findings related to policy and abortion coercion. |
| **Research/policy recommendations** | Study author-identified research/policy recommendations |
| **Gaps/Areas of Further Research** | Existing research gaps and areas of further research needed, as identified by study authors |

1 Adapted from Pollock *et al. 2023*
